# Combining Procedural and Behavioral Treatments for Chronic Low Back Pain: A Pilot Feasibility Randomized Controlled Trial

**DOI:** 10.1101/2023.06.02.23290392

**Authors:** Adrienne D. Tanus, Isuta Nishio, Rhonda Williams, Janna Friedly, Bosco Soares, Derek Anderson, Jennifer Bambara, Timothy Dawson, Amy Hsu, Peggy Y. Kim, Daniel Krashin, Larissa Del Piero, Anna Korpak, Andrew Timmons, Pradeep Suri

**Author notes:** Corresponding author: Adrienne D. Tanus, MPH, VA Puget Sound Health Care System, S-152-ERIC, 1660 S. Columbian Way, Seattle, WA, 98108.

## Abstract

Individual treatments for chronic low back pain (CLBP) have small magnitude effects. Combining different types of treatments may produce larger effects. This study used a 2×2 factorial randomized controlled trial (RCT) design to combine procedural and behavioral treatments for CLBP. The study aims were to: (1) assess feasibility of conducting a factorial RCT of these treatments; and (2) estimate individual and combined treatment effects of (a) lumbar radiofrequency ablation (LRFA) of the dorsal ramus medial branch nerves (vs. a simulated LRFA control procedure) and (b) Activity Tracker-Informed Video-Enabled Cognitive Behavioral Therapy program for CLBP (AcTIVE-CBT) (vs. an educational control treatment) on back-related disability at 3 months post-randomization. Participants (n=13) were randomized in a 1:1:1:1 ratio. Feasibility goals included an enrollment proportion ≥30%, a randomization proportion ≥80%, and a ≥80% proportion of randomized participants completing the 3-month Roland-Morris Disability Questionnaire (RMDQ) primary outcome endpoint. An intent-to-treat analysis was used. The enrollment proportion was 62%, the randomization proportion was 81%, and all randomized participants completed the primary outcome. Though not statistically significant, there was a beneficial, moderate-magnitude effect of LRFA vs. control on 3-month RMDQ (−3.25 RMDQ points; 95% CI: -10.18, 3.67). There was a significant, beneficial, large-magnitude effect of AcTIVE-CBT vs. control (−6.29, 95% CI: -10.97, -1.60). Though not statistically significant, there was a beneficial, large effect of LRFA+AcTIVE-CBT vs. control (−8.37; 95% CI: -21.47, 4.74). We conclude that it is feasible to conduct an RCT combining procedural and behavioral treatments for CLBP.

ClinicalTrials.gov Registration: https://clinicaltrials.gov/ct2/show/NCT03520387

## Introduction

Low back pain is the leading contributor to years lived with disability and health care costs in the United States (US).[13; 28] This is driven mainly by chronic low back pain (CLBP; pain persisting for ≥3 months).[20] Most treatments for CLBP have only small magnitude effects compared with control treatments, i.e., roughly 5-10% improvements in functional limitations and disability in randomized controlled trials (RCTs).[11]

Combining different treatments may be one way to increase the magnitude of CLBP treatment effects.[29] An important, unanswered question is “Which interventions should be combined?”. In considering this question, we applied the theoretical framework of the Nagi disablement model, in which pain is considered an impairment on the pathway to disability (**Figure 1**). We reasoned that attaining large treatment effects on disability for people with CLBP would require combinations of treatments that address different stages on the disablement pathway. Interventional procedural treatments for CLBP target pathoanatomic “pain generators” in the spine, potentially improving disability by affecting the underlying structures which cause CLBP, or by decreasing pain intensity. In contrast, behavioral treatments for CLBP such as cognitive behavioral therapy (CBT) aim to improve functional limitations and disability by modifying how people think about pain or how pain affects their behavior. Therefore, combining procedural and behavioral treatments for CLBP may provide more opportunities for improving functional outcomes than would a single treatment alone. However, no prior RCTs have examined whether interventional procedural treatments and behavioral treatments for people with CLBP can feasibly be combined in an RCT. A key concern is whether people who have sought out interventional pain procedures targeting specific anatomic pain generators - an approach grounded in a biomedical model of pain - would be willing to be randomized to a behavioral pain treatment, a fundamentally different approach that primarily targets the psychosocial components of chronic pain.

**Figure 1.**
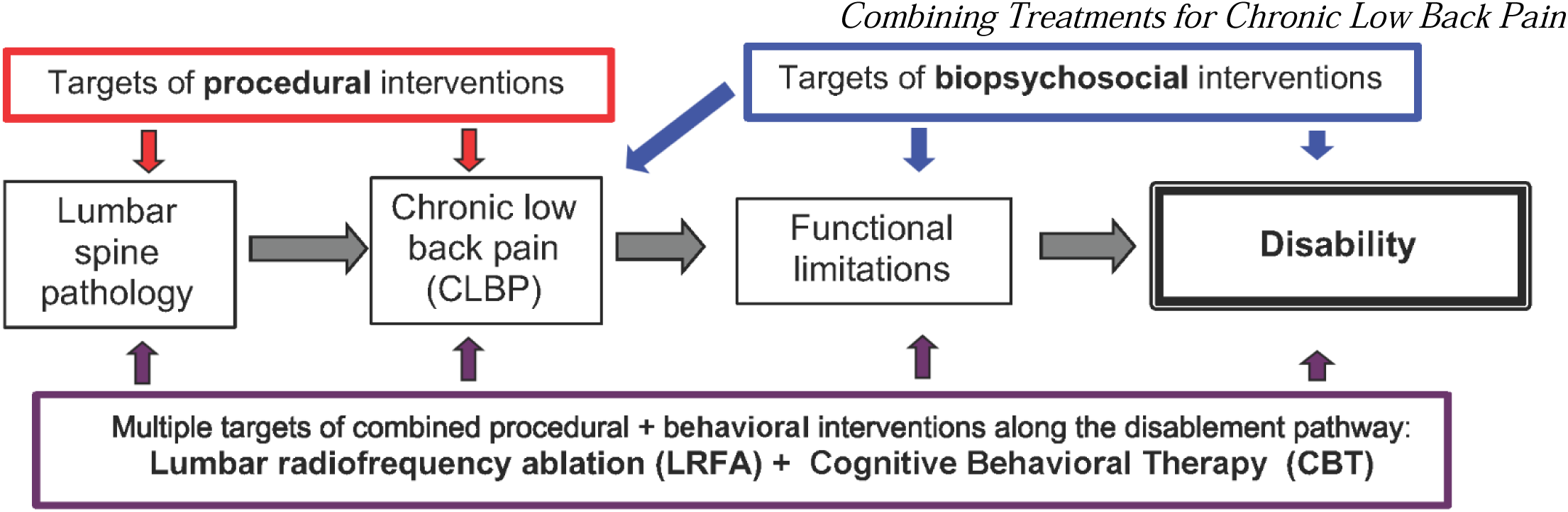
Conceptual model of potential targets for interventions in chronic low back pain (CLBP).

We conducted a pilot feasibility RCT to examine whether an interventional procedural treatment for CLBP, lumbar radiofrequency ablation (LRFA), could be combined with a video telehealth-based behavioral treatment for pain, cognitive behavioral therapy (CBT). LRFA is a minimally invasive procedure that can result in large-magnitude improvements in properly selected individuals with CLBP.[14] However, there is conflicting evidence regarding its efficacy,[22; 24] and no treatment effect was found in the largest RCT conducted.[19] In contrast, CBT is a widely accepted treatment for CLBP with demonstrated effectiveness,[8; 9] but typically modest magnitude effects.[37] In-person CBT is not widely available in the US[16] and incurs substantial travel burdens for patients, which motivated this examination of video-based CBT. The first study aim was to evaluate the feasibility of conducting a factorial RCT of LRFA and video-based CBT, each compared to an active control condition. The second aim was to estimate the individual and combined treatment effects of (1) LRFA (vs. a simulated LRFA control procedure) and (2) video-based CBT (vs. an educational control treatment) on back-related disability at 3 months post-randomization.

## Methods

### Study design

This was a 2 x 2 factorial pilot RCT (Figure 2). Although sometimes conceptualized as a study design that allows examination of interactions between treatment effects, factorial trials are typically underpowered for evaluating interactions.[26] Instead, the factorial RCT is more accurately viewed as an efficient trial design that permits the study of two interventions at the same time in the same study sample, with each intervention compared to control treatments.[26] Eligible participants were randomized to receive LRFA vs. simulated LRFA with corticosteroid injections to the lumbar medial branch nerves. Simultaneously, the same participants were randomized to receive an Activity Tracker-Informed Video-Enabled Cognitive Behavioral Therapy program for CLBP (AcTIVE-CBT) delivered using personal computers (PC) or tablets vs. a brief program of telephone-based self-directed CBT and education (TBSCE). Participants were randomized in a 1:1:1:1 ratio to each of the 4 cells of the contingency table (Figure 2). All research processes were approved by the VA Puget Sound Institutional Review Board (MIRB01676). The study was registered at clinicaltrials.gov (https://clinicaltrials.gov/ct2/show/NCT03520387) prior to the start of recruitment; updates to the trial registry were required by the sponsor every 6 months until study completion.

**Figure 2.**
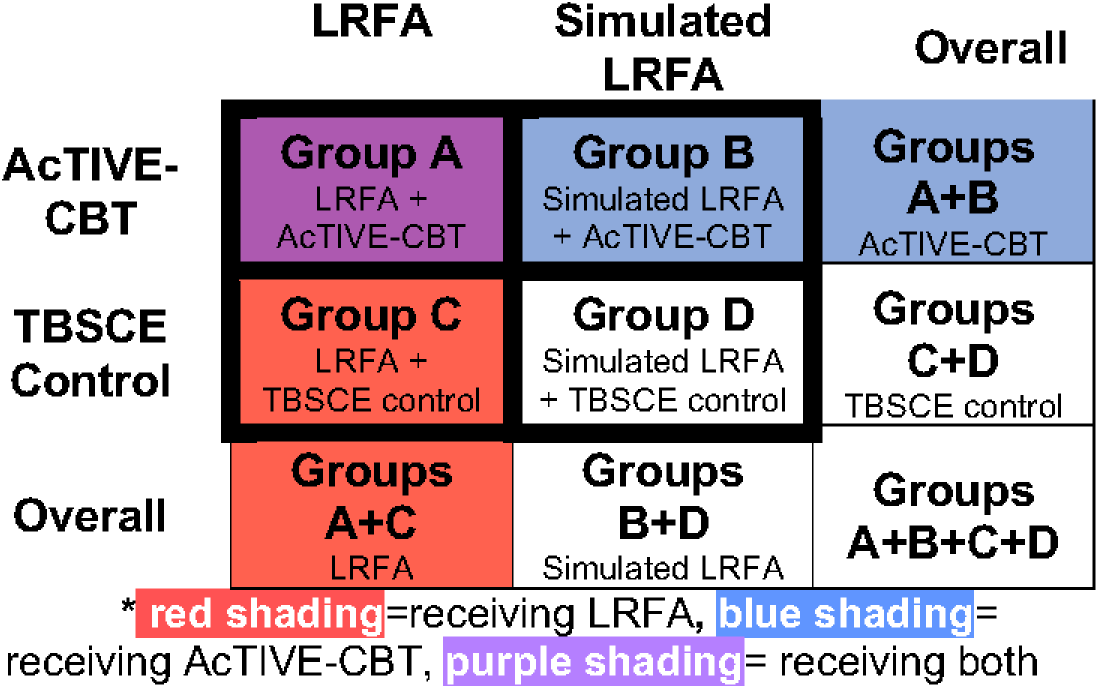
The 2 x 2 factorial trial study design*

### Participants

LRFA can be used for people with CLBP attributed to the lumbar facet joints, defined by pain relief to local anesthetic blocks along the dorsal ramus medial branch nerves.[3; 15] LRFA applies a thermal lesion to these nerves, temporarily reducing or eliminating CLBP. Because lumbar facet joint-mediated pain cannot be diagnosed by clinical characteristics or imaging, in typical clinical care, possible candidates for LRFA are selected according to whether or not they have low back pain relief (“positive responses”) with anesthetic blocks of the medial branch nerves (“MBBs”), which innervate the lumbar facet joints.[15] The target population for this study consisted of adult patients with CLBP seeking care at a single center, the Veterans Affairs (VA) Puget Sound Health Care System, and receiving lumbar medial branch blocks (MBBs) as part of usual clinical screening for LRFA eligibility. Clinical screening involved a 1^st^ set of MBBs, followed by a 2^nd^ set of MBBs (performed only for those patients with positive responses to the 1^st^ set of MBBs), followed by LRFA (performed only for patients with positive responses to the 2^nd^ set of MBBs). Potential study participants were approached during this routine clinical process for determining LRFA eligibility.

Eligible participants were Veterans of the Armed Forces receiving care at VAPSHCS who had: (1) low back pain of ≥3 months duration; (2) low back pain intensity ≥4 on the numerical rating scale (NRS); (3) previously received 1^st^ line rehabilitative treatments, such as physical therapy or spinal manipulation; (4) the ability to provide informed consent and complete assessments; (5) access to a personal electronic device with Internet access; (6) ”positive responses” to 2 separate sets of lumbar medial branch blocks (low volume anesthetic blocks of the medial branches of the dorsal rami using 0.5cc or less of lidocaine or bupivacaine), with at least 50% pain improvement in the low back pain intensity NRS; and (7) were a candidate for unilateral or bilateral LRFA between the L1 and S1 spinal levels. Exclusion criteria were: (1) a diagnosis of lumbosacral radiculopathy, symptomatic lumbar spinal stenosis (neurogenic claudication), spinal instability, infection, malignancy, or fracture; (2) pregnant females, prisoners, or cognitively impaired; (3) prior lumbar RFA; (4) prior lumbar spine surgery involving the levels where LRFA was to be performed within the past 2 years; (5) ever having received lumbar fusion involving the levels where LRFA was to be performed; (6) prior CBT for chronic pain; (7) primary psychotic or major thought disorder, any active suicidal/homicidal ideation, or unstable or severe psychiatric/behavioral conditions; (8) hospitalization for psychiatric reasons other than suicidal ideation, homicidal ideation, and/or PTSD, in the past 5 years; (9) cognitive limitations as assessed by a validated 6-item cognitive screener;[6] or (10) severe active medical comorbidities limiting study participation. As a component of this feasibility study, some inclusion and exclusion criteria were modified during the course of the study (Supplemental Table S1). Modifications to study criteria generally involved liberalizing study-related restrictions, so as to have the target population more closely represent the typical patients being evaluated for LRFA in usual clinical care at our facility, while increasing the recruitment rate.

### Recruitment

Supplemental Figure S1 shows participant flow through study processes. Eligible participants were recruited on a rolling basis from November 2018 to March 2020, when recruitment was terminated due to the COVID-19 pandemic. Study staff reviewed electronic health records of all patients scheduled for upcoming lumbar MBBs to identify potential participants. Clinical providers were alerted to potentially eligible study candidates; providers then introduced the study and inquired about patients’ willingness to talk to research staff about the study. Interested patients were assessed for eligibility screening by research staff and received information about the study after their 1^st^ set of MBBs. Patients who met preliminary eligibility criteria were again approached after their 2^nd^ set of MBBs, at which time eligibility criteria were reassessed and eligible patients were offered informed consent. Following enrollment into the study, participants were given a baseline assessment and received a Fitbit Zip unit (Fitbit Inc., San Francisco, CA) to be worn for the entire study participation period. A run-in period was used between enrollment and randomization to exclude participants who could not be contacted by research staff or were unable to use the Fitbit units, to ensure that randomized participants were capable of completing follow-up assessments. On the day of randomization and prior to randomization, participants were re-assessed to ensure that eligibility criteria were met; those ineligible at these reassessments were not randomized.

### Randomization and Blinding

Computer-generated permuted-block randomization with a 1:1:1:1 ratio (Figure 2), stratified by MBB block responses (50-79% pain relief vs. ≥80% pain relief), was used to achieve roughly balanced groups. Participant group allocations were generated by a data team member who did not conduct any patient-facing procedures, and allocations were concealed from research staff in sealed, numbered envelopes kept in a secure, locked cabinet. Envelopes were assigned to participants in sequence, stratified by MBB block responses. Envelopes were only opened by a non-blinded research staff member immediately prior to the lumbar procedure, and a non-blinded research interventional pain physician, after confirming that the participant remained eligible for randomization at the time of the scheduled spine procedure. Group allocations were not divulged to other research staff members, including those involved in study assessments, who remained blinded to allocation until after the primary study end point at 3 months post-randomization.

Eligible participants received screening diagnostic lumbar MBBs and all aspects of the LRFA procedures prior to medial branch lesioning or simulated lesioning by a blinded “treating interventional pain physician” who was the primary pain provider for the participant. The treating interventional pain physician conducted needle and electrode placements for the LRFA or simulated LRFA procedure, as well as motor and sensory stimulation testing of the medial branch nerves to confirm placement and safety. Immediately prior to medial branch nerve lesioning or simulated lesioning, the blinded treating interventional pain physician left the procedure room, and lesioning or simulated lesioning was conducted by the non-blinded research interventional pain physician. This process permitted blinding of the treating interventional pain physician to the procedural treatment received on the day of the procedure. The treating interventional pain physician was instructed to not attempt to discover the actual LRFA treatment status (LRFA vs. simulated LRFA) of their patient. All interventional care following the day of the procedure was conducted by pain clinic staff as per usual clinical care, with oversight of the blinded treating interventional pain physician when needed, as if all participants had received LRFA. In this way, the treating interventional pain physician remained blinded to the procedural treatment received until after the primary end point at 3 months post-randomization. Similarly, the participant and the research staff conducting study assessments were blinded to the procedural treatment received until after the primary end point at 3 months post-randomization. Thus, a triple-blind design was achieved for procedural treatments.

Treating psychologists could not be blinded to the behavioral treatment provided (AcTIVE-CBT vs. TBSCE). Participants were aware of which behavioral treatment they were receiving, but were not informed which of the two treatments was considered the study “intervention” of interest. The research staff conducting study assessments were blinded to the behavioral treatment received until after the primary end point at 3 months post-randomization. Thus, a single-blind design was achieved for behavioral treatments.

### Study Interventions (Procedural)

All procedural treatments were delivered by physicians who were board-certified in pain medicine. All interventionists (both procedural and behavioral) in this study were licensed independent practitioners on staff at VAPSHCS, providing the interventions within the context of clinical care.

#### LRFA procedural technique

Participating interventional physicians were instructed to follow LRFA technical recommendations with regards to parallel electrode placement and the use of two radiofrequency lesions per medial branch as per the Spinal Intervention Society (SIS) guidelines, 2nd edition.[4] This involved placement of an 18-gauge radiofrequency electrode along the course of each medial branch targeted, using a fluoroscopic declined view approach with cross-table obliquity as needed to achieve electrode placement parallel to the medial branch. Although not part of the SIS guidelines, all interventional physicians used sensory and motor stimulation testing as part of their usual clinical practice. Once each electrode was positioned, sensory stimulation was performed at 50 Hz up to 1 millivolt until the participant reported axial and no extremity sensation to confirm appropriate location of the electrode. Motor stimulation was performed at 2 Hz up to 1 volt to confirm that there was no motor stimulation of the ventral nerve root causing muscle contraction. If there was motor stimulation of the ventral nerve root, the electrode was repositioned, and the stimulation sequence was repeated until the correct electrode position was confirmed. Once the electrode was in correct position and stimulation testing completed, a lesion was generated by raising the temperature at 1°C per second, from 37°C to an operating temperature between 80°C and 85°C, which was maintained for 90 seconds. The electrode was then repositioned by withdrawing or repositioning parallel to the first ablation site, or by rotating the electrode, and a second lesion was made. If at any time during the raising of the temperature, or during the coagulation, the participant reported adverse sensations, the generator was immediately turned off and the sensation evaluated. The decision of whether to use post-LRFA corticosteroid medial branch injections to mitigate post-LRFA neuritis and the manner in which this was done (including corticosteroid type and dose) was left to the usual clinical practices of each interventional physician; all physicians routinely used post-LRFA corticosteroid medial branch injections. This process was then repeated for each medial branch nerve that was targeted (2 or more medial branches per LRFA procedure).

#### Simulated LRFA with targeted steroid injection procedural technique

The simulated LRFA control was performed in an identical fashion to LRFA as above, except that after electrode positioning and sensory/motor testing, a neurodestructive lesion was <u>not</u> made. Instead, a pre-recorded audio recording of the procedure was played by a clinical assistant who was out of view of the participant, in order to simulate the beeping and other sounds of the radiofrequency generator machine and to ensure the appropriate length of the simulated procedure. At each medial branch site, the electrode remained in place for the full 90 seconds that lesioning would normally require, but without heat application. The electrode was then repositioned, and a second simulated lesion of duration 90 seconds was applied, along with the audio recording. This process was then repeated for each medial branch nerve that was targeted (2 or more medial branches per simulated LRFA procedure). Targeted corticosteroid injections consisted of a total injectate quantity per participant not to exceed the equivalent of 80 mg triamcinolone, divided equally among the medial branch sites targeted.

### Study Interventions (Behavioral)

All behavioral treatments were delivered by licensed clinical psychologists with expertise in rehabilitation and cognitive-behavioral therapy and its application to the treatment of chronic pain. Participants receiving a given treatment (AcTIVE CBT or TBSCE) generally received the same information according to a protocol (full descriptions of each treatment provided in Supplemental Table S2); however, psychologists were permitted to tailor sessions as needed at their discretion to ensure participant understanding of the content, and to provide individualized support based on personalized goals, barriers, and abilities.

#### Video-based CBT (AcTIVE-CBT)

AcTIVE-CBT involved eight 60-minute treatment sessions spaced over 3 months, delivered via 1:1 clinical video telehealth (CVT) to participants in their homes. The AcTIVE-CBT content closely emulated that used in the Mind-body Approaches to Pain (MAP) trial,[8; 9] with modifications to fit the CVT context and shorter (60-minute) treatment sessions. Psychologists were encouraged to incorporate information provided by the Fitbit Zip units into treatment sessions, by referring to Fitbit output during the initial evaluation and each treatment session, including reminders and tips about Fitbit use, evaluating step count homework, and tracking overall progress towards goals.

Psychologists providing video-based CBT received reports on a participant’s Fitbit output and compliance in advance of each treatment session. Psychologists attempted to schedule weekly sessions at consistent days and times, to facilitate routine and attendance, and to ensure adequate practice time for participants between sessions. Sessions began with a review of the Fitbit-related output provided by research staff, to gauge activity since the prior session. Sessions typically included activity plans, expectations, and goals for the next session and for the long-term. Some differences between AcTIVE-CBT and the content of the MAP trial[8; 9] included a greater focus on encouraging walking and physical activity, use of the Fitbit Zip units to provide objective assessments of physical activity, and slightly greater emphasis on understanding flare-ups and relapse prevention. AcTIVE-CBT included the same audio content developed in the MAP trial (e.g., guided breathing and relaxation scripts), which participants were instructed to use for home practice. This content was provided to participants in CD format and was also accessible online. The AcTIVE-CBT treatment manual is provided as a Supplemental File.

#### Telephone-based self-directed CBT and education (TBSCE)

The TBSCE control treatment was a self-directed, bibliotherapy-based intervention, with some support from a psychologist in a structured introductory session at the beginning and a booster session at 3 months post-randomization. These sessions were provided by the same clinical psychologists who provided AcTIVE-CBT. TBSCE involved an initial 60-minute telephone education session by a psychologist including education on CBT principles; the provision of an educational book for CBT self-management and orientation to the book;[7] and a plan for weekly reading and homework using the book. In addition, the same psychologist who conducted the initial telephone education session contacted participants at the midpoint of the 3-month follow-up period to provide support, reinforce key concepts as needed, encourage continued participant-directed use of the plan for weekly reading and workbook use, and answer questions. Weekly workbook homework included: 1) education on chronic pain, 2) theories of pain and diaphragmatic breathing, 3) progressive muscle relaxation and visual imagery, 4) automatic thoughts and pain, 5) cognitive restructuring, 6) stress management, 7) time-based pacing, 8) pleasant activity scheduling, 9) anger management, 10) sleep hygiene, and 11) relapse prevention and flare-up planning. Participants in the TBSCE arm also received the Fitbit Zip units, but education and treatment were not specifically structured around use of the Fitbit.

### Study Measures

Study assessments up to and including the day of randomization were conducted by in-person interviews. Subsequent data collection was done by telephone interview, unless in-person assessments were requested by the participant. Fitbits were worn by participants throughout study participation. Fitbit data was synced and downloaded on a weekly basis. Post-randomization study assessments were conducted monthly, and the main study outcomes were assessed at 3 months post-randomization as pre-specified in the trial registration.

#### Feasibility outcomes

As some aspects of study feasibility were unclear prior to the start of the study, given the factorial design and randomization to both procedural and behavioral treatments, specific feasibility outcomes and targets were not formally specified until after recruitment was initiated. Accordingly, only effectiveness outcomes (and not feasibility outcomes) were specified in the trial registration. Quantitative targets used to judge the feasibility of a future trial included (1) a proportion of those enrolled from among those meeting all study criteria prior to LRFA ≥30% (the recruitment proportion achieved in a prior sham-controlled RCT by our group);^33^ (2) a proportion of those randomized (from among those enrolled) of ≥80%, and (3) a proportion of randomized participants completing the 3-month RMDQ primary endpoint of ≥80%.

#### Effectiveness outcomes

##### Primary Outcome (the Roland-Morris Disability Questionnaire [RMDQ])

The RMDQ score at 3 months post-randomization was the primary study endpoint effectiveness outcome. The RMDQ is a validated and reliable back pain-specific functional status questionnaire adapted from the Sickness Impact Profile.[31] The RMDQ consists of 24 yes/no items which represent common limitations in daily activities experienced by those with low back pain. A single unweighted score is derived by summing the 24 items, with scores ranging from 0 (no disability) to 24 (maximum disability) and higher scores indicating worse function. The RMDQ was designed for paper administration and is also well-suited for telephone use.[31] It is recommended as a core outcome measure by the US National Institutes of Health (NIH) Task Force on Research Standards for Chronic Low Back Pain.[12]

##### Secondary Outcomes

Participant activity was tracked pre- and post-randomization to assess change in step counts. Average daily step counts were measured using the Fitbit Zip units, which have demonstrated reliability and validity as compared to actual observed step counts or reference standard activity monitors.[30] Baseline average step counts were calculated as the average daily step counts over the day of the baseline survey and the two prior days. Average step counts at 3-month follow-up were calculated as the average daily steps in the 2 weeks prior to survey, among days on which participants wore the Fitbit. Change in step counts was calculated as the difference between average step counts at 3-month follow-up and baseline. CLBP intensity was measured using a 0-10 Numeric Rating Scale (NRS) of pain over the past 24 hours, in which “0” reflects no pain and “10” reflects the “worst pain imaginable”. The pain NRS is valid, reliable, and sensitive to detecting change in pain intensity after treatment.[18] Quality of life was measured using the mental health and physical health subscales of the PROMIS Short Form 10, which has been recommended as a core outcome to be used in clinical trials.[10] Average morphine equivalent daily dose was calculated based on participant self-report of opioids used in the past 72 hours. At treatment completion only, participants rated the global perceived effect of treatment using a 7-point Likert scale (completely recovered, much improved, improved, not changed, worsened, much worsened, become worse than ever) and satisfaction with treatment using a 5-point Likert scale (very satisfied, somewhat satisfied, neutral, somewhat dissatisfied, very dissatisfied). Participants also completed a novel measure of pain intensity, the Numeric Rating Scale of Underlying Pain without concurrent Analgesic use (the NRS-UP_(A)_). Participants who reported any analgesic use in the past 72 hours were also asked to make a second report of what they expected their NRS pain rating for CLBP in the past 24 hours *would have been* if they were not taking analgesics, which we refer to as the NRS-UP_(A)_ item. The NRS-UP_(A)_ is defined as the value of the NRS-UP_(A)_ item among participants who reported any analgesic use, and the value of the conventional NRS of CLBP intensity among participants who were not taking analgesics. The NRS-UP_(A)_ was pre-specified as a secondary outcome, but the measure is still under development and has not yet been validated. Participants reported average minutes spent engaging in moderate and heavy physical activity each week using items from the Behavioral Risk Factors Surveillance System (BRFSS),[32; 38] which were used to calculate “moderate intensity-equivalent minutes” per week using an established method.[5]

### Blinding

Although no formal assessment of blinding was made post-procedure on the day of randomization, blinding was evaluated monthly thereafter. The primary time point for assessing blinding was 1 month post-procedure, the earliest time at which blinding was assessed, as it was thought that assessing blinding later in follow-up may lead to situations where a participant’s perception of their response to treatment would influence their view of whether they had received LRFA vs. simulated LRFA. Participants were asked to guess whether they received LRFA, simulated LRFA, or whether they had “no idea”. Those who guessed LRFA or simulated LRFA were also asked whether they based their guess on a particular clue. The blinding index described by Bang et al. was used to evaluate the statistical significance of participants’ tendency to guess the treatment they received (blinding).[2]

### Adverse events

Adverse events (AEs) were defined as any untoward occurrence that presented during follow-up, regardless of whether or not the event could have had a causal relationship with the study interventions. Adverse events were recorded at the time of identification and classified by severity, relationship to study procedures, and whether the AE was an anticipated event following the study interventions. All AE outcomes were followed until resolution. Serious adverse events (SAEs) were defined as AEs that resulted in death; a life-threatening problem, incapacity, disability; hospitalization; or medical intervention to prevent impairment or damage. Two methods were used to monitor for AEs: active identification through participant reporting at the time of each monthly scheduled assessment, and passive identification through review of the medical record on a monthly basis. AE analyses were specified *a priori*. AEs and the results of AE analyses were reviewed by an independent Data Monitoring Committee at the midpoint and conclusion of data collection.

### Sample size

As this was a feasibility study, one goal of which was to estimate the annual recruitment capacity at our institution, we did not conduct formal sample size calculations. Based on the 2-year duration of funding, expected time for study start-up, and estimated number of LRFA procedures conducted at our facility, recruitment goals were to randomize 20 Veterans over 12-18 months of recruitment. Assuming that as many as 1 out of every 3 Veterans enrolled would withdraw between enrolment and randomization, we planned to enroll up to 30 Veterans.

### Statistical Analyses

As appropriate to the 2 x 2 factorial design,[26] first, we compared participants randomized to LRFA and simulated LRFA. Next, we compared participants randomized to AcTIVE-CBT and TBSCE. Feasibility outcomes were evaluated based on whether the proportions attained achieved planned targets. Effectiveness outcomes were between-group differences evaluated with an intent-to-treat strategy, using linear regression models with the outcome measure at 3 months post-randomization as the dependent variable, an indicator of treatment group as the independent variable, and adjustment for age and baseline value of the outcome measure where a baseline value was relevant; for example, the outcome of participant satisfaction with treatment had no baseline value (because treatment occurs after the baseline) to include in the model. The primary outcome was RMDQ scores at 3 months post-randomization. We fit a separate regression model for each outcome and calculated 95% confidence intervals (CIs) for treatment coefficients; statistical significance was defined as 95% CIs excluding a null effect (0 for continuous outcomes). Interpretation was focused on the effect size point estimates, rather than statistical significance, in keeping with the limited sample size of this feasibility study. Negative values of the treatment effect indicated better scores on the outcome variable with the active treatment for models examining RMDQ (the primary outcome), the NRS, morphine equivalent daily dose, and the NRS-UP_(A)_. Positive values of the treatment effect indicated better scores on the outcome variable with the active treatment for models examining change in step counts, the PROMIS mental health and physical health subscales, global perceived effect, satisfaction, and BRFSS moderate intensity-equivalent minutes. First, we evaluated model results for the RMDQ to examine for large-magnitude interactions on the additive scale, by qualitatively comparing the model-estimated treatment effects for both active treatments combined (patients receiving LRFA + AcTIVE-CBT [Figure 2, Group D] vs. simulated LRFA + TBSCE [Figure 2 Group D] ; Figure 2, Group A) with the “main effects” of LRFA (patients receiving LRFA [Figure 2, Groups B+D] vs. simulated LRFA + TBSCE [Groups A+C]) and AcTIVE-CBT (patients receiving AcTIVE-CBT [Figure 2, Groups C+D] vs. TBSCE [Group A+B]). If model-estimated treatment effect point estimates for both active treatments combined did not show large differences from the sum of each active treatment main effect point estimate, no interactions were assumed in subsequent analyses. We then repeated this process for the secondary outcomes. Binary “responder” outcomes for the RMDQ and NRS were not analyzed due to the very small sample size and consequent imprecision of binary outcomes.[1]. We calculated the Bang blinding index, which produces an estimate which ranges from −1 to 1, with −1 indicating that all participants guessed the incorrect treatment, 0 indicating that participants randomly guessed their study-group assignments, and 1 indicating that all participants correctly guessed their study-group assignments.[2] No interim analyses or stopping guidelines were planned, in keeping with the nature of this study as a pilot.

## Results

### Study Participants

A total of 109 patients were screened between November 2018 and March 2020 (**Figure 3**). Of these, 26 patients met eligibility criteria for recruitment. Major reasons for not meeting eligibility criteria included having undergone prior LRFA or lumbar spine surgery, receiving <50% relief in pain after MBBs or experiencing extended pain relief after MBBs, and having an NRS score of ≤3. Of 26 patients who met study criteria after their 2nd set of MBBs, 16 (62%) were enrolled in the study. Of these 16 participants, two failed to meet study criteria prior to the date of randomization due to extended pain relief or lack of ≥50% pain relief with the 2nd set of MBBs, and one declined to participate prior to the date of randomization, leaving thirteen out of 16 enrolled participants (81%) who were randomized. All participants who were randomized received their allocated procedural treatment; however, 1 participant who was randomized to LRFA and who had received bilateral MBBs for bilateral CLBP (pain on both sides of the low back) received LRFA on only 1 side of the low back. All participants who were randomized received their allocated behavioral treatment: the 7 participants randomized to AcTIVE-CBT received all eight planned treatment sessions; and of the 6 participants randomized to TBSCE, 5 received both planned treatment sessions and 1 received only one session. All participants completed the main follow-up at 3-months post-randomization.

**Figure 3.**
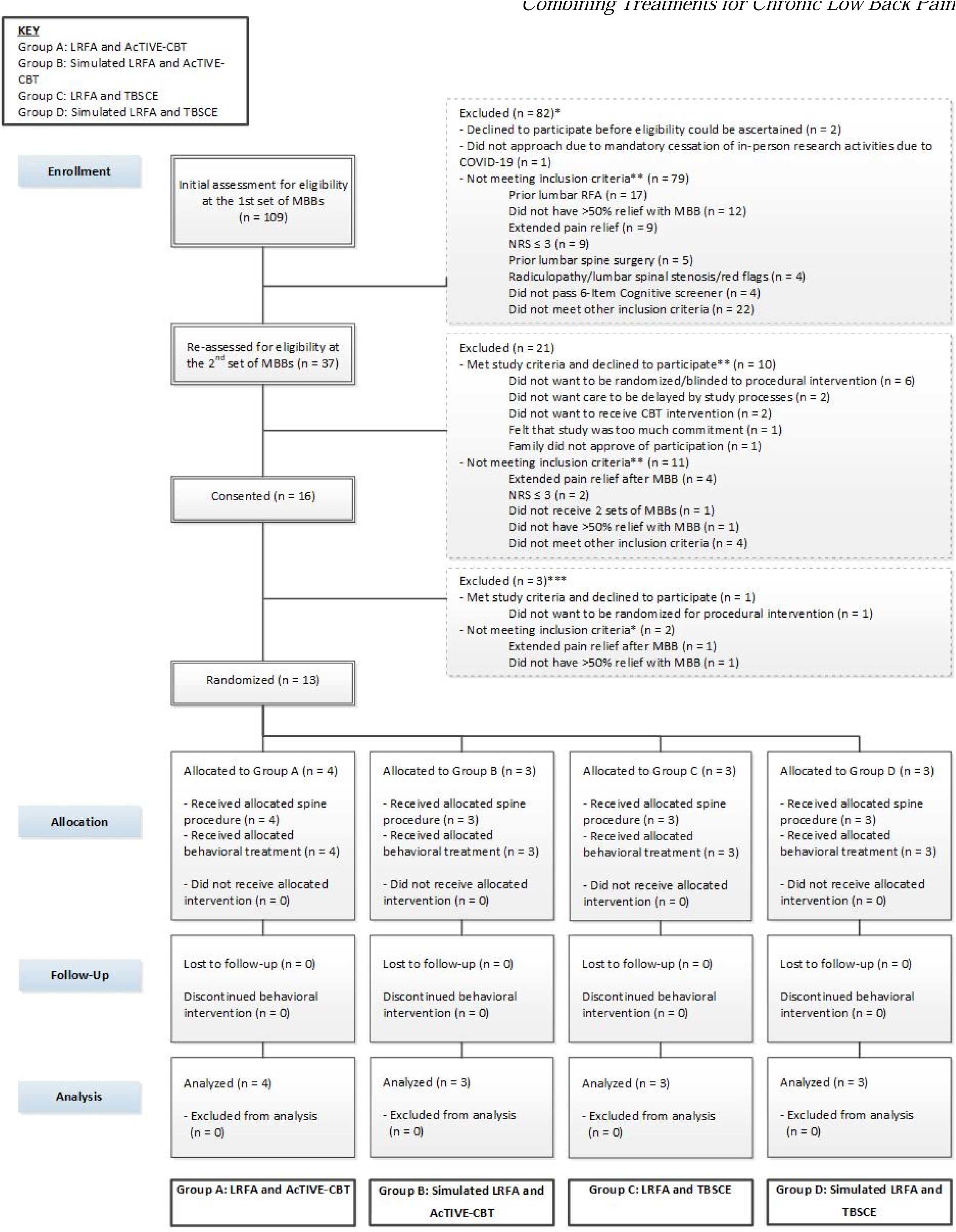
Enrollment, randomization, and follow-up. *Number of potential participants screened at time of 1^st^ MBBs minus those excluded before re-assessment may not add up to the total re-assessed, as the initial assessment for some potential participants may take place at the time of the 2^nd^ set of MBBs **Numbers may not add up to the total as potential participants may not meet multiple exclusion criteria or may decline participation for more than one reason ***Twenty-six potential participants met study criteria at the time of consent, including 16 who were consented and 10 who declined to participate

All 13 randomized participants self-identified as male and the mean age of participants was 58 years. The majority of participants were white (69%), and 23% of participants were Hispanic or Latino (Table 1). Participants receiving LRFA and simulated LRFA were comparable with regards to many characteristics, but those receiving LRFA were generally older (62 vs. 54 years) and had higher levels of baseline disability on the RMDQ (15 vs. 12 RMDQ points) (Table 1). Participants receiving AcTIVE-CBT and TBSCE were comparable in regard to many characteristics, but those receiving AcTIVE-CBT were more likely to be Latino or Hispanic (43% vs. 0). There were minor differences in baseline disability between those who received AcTIVE-CBT and TBSCE (13 vs 15 RMDQ points) (Table 1). Participants receiving LRFA/AcTIVE-CBT and simulated LRFA/TBSCE were generally comparable, but those receiving LRFA/AcTIVE-CBT were older (59 vs. 49 years) and were more likely to be Latino or Hispanic (50% vs. 0) (Supplemental Table S1).

**Table 1.**
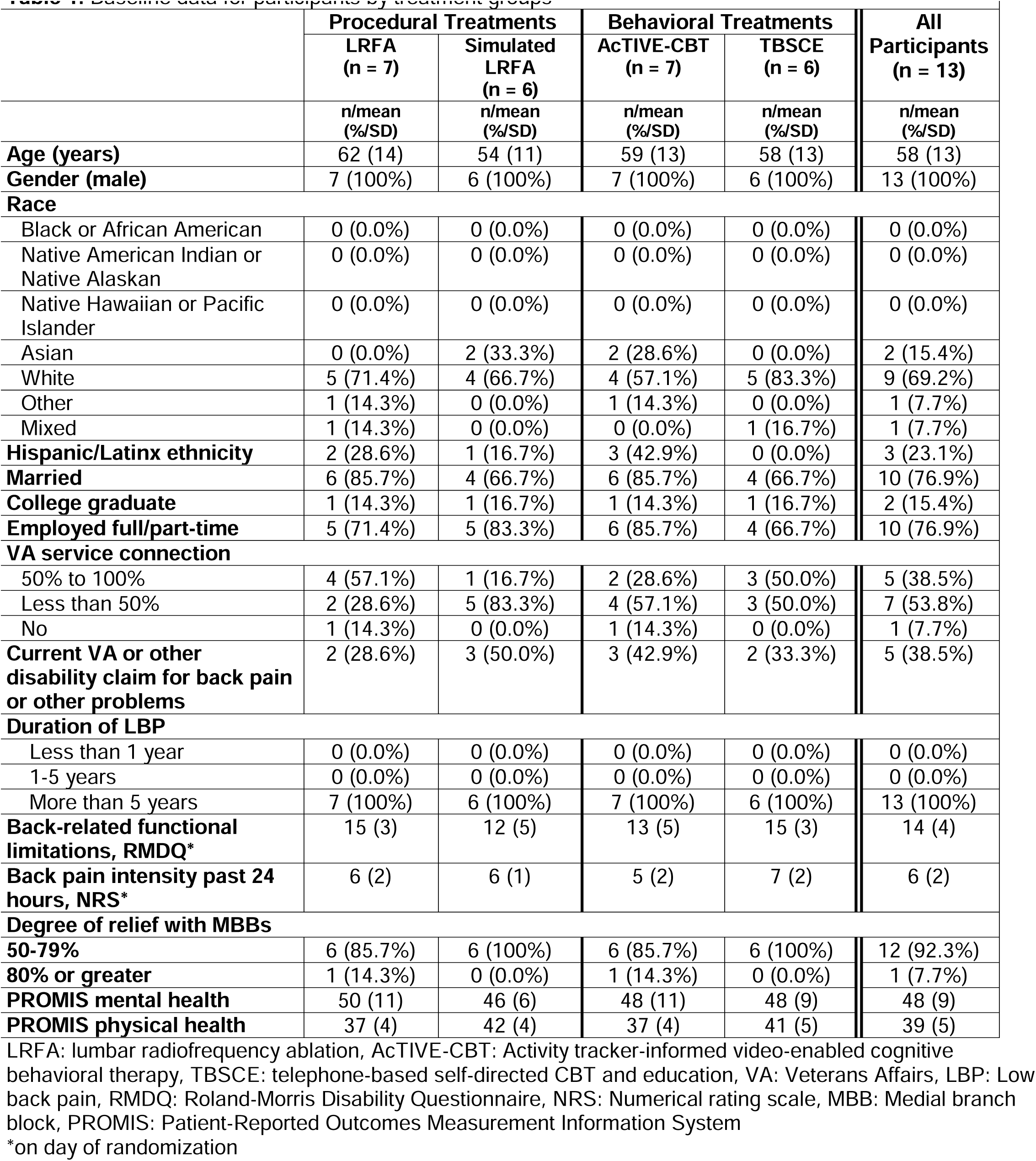
Baseline data for participants by treatment groups

### Qualitative evaluation of interactions between treatment effects

Descriptive evaluation of plots of RMDQ scores showed post-randomization improvements in the active treatment groups as compared to the control groups, with substantially larger post-randomization improvements for participants in the AcTIVE-CBT vs. TBSCE and LRFA + AcTIVE-CBT vs. simulated LRFA + TBSCE groups, as compared to participants in the LRFA vs. simulated LRFA group (Supplemental Figures S2-S4). The largest between-group difference in post-randomization change occurred between the day of randomization and 1 month post-randomization.

Treatment effect point estimates from models examining RMDQ outcomes showed no suggestion of large-magnitude interactions on the additive scale between the effects of LRFA (vs. simulated LRFA) and AcTIVE-CBT (vs. TBSCE) treatments for the primary RMDQ outcome (Table 2). This is demonstrated by the treatment effect point estimate of LRFA vs. simulated LRFA (−3.3 RMDQ points) and the treatment effect point estimate of AcTIVE-CBT vs. TBSCE (−6.3 RMDQ points), which add up to a total (−9.6 RMDQ points) that is only slightly larger than the estimated treatment effect of LRFA+AcTIVE-CBT vs. simulated LRFA vs. TBSCE (−8.4 treatment effect points). This suggested approximately additive treatment effects of LRFA and AcTIVE-CBT; accordingly, no interactions on the additive scale were assumed in subsequent analyses.

**Table 2.** Multivariate associations between variables and outcomes*

| **Table 2. Multivariate associations between variables and outcomes\*** |  |  |  |
| --- | --- | --- | --- |
|  | **LRFA vs.**<br>**simulated LRFA**<br>**(n =13)** | **AcTIVE-CBT vs.**<br>**TBSCE**<br>**(n = 13)** | **LRFA + AcTIVE-CBT**<br>**vs. simulated LRFA +**<br>**TBSCE**<br>**(n = 13)** |
|  | **$\beta$ (95% CI)** | **$\beta$ (95% CI)** | **$\beta$ (95% CI)** |
|  | **Primary outcome** |  |  |
| RMDQ<sup>a</sup> | -3.3 (-10.2, 3.7) | -6.3 (-11.0, -1.6) | -8.4 (-21.5, 4.7) |
|  | **Secondary outcomes** |  |  |
| Change in step counts<sup>bc</sup> | -6059 (-12467, 349) | 2082 (-4667, 8830) | -2824 (-10622, 4972) |
| NRS<sup>a</sup> | -1.0 (-5.2, 3.1) | -0.3 (-4.2, 3.6) | -0.8 (-13.4, 11.7) |
| PROMIS GH<sup>b</sup> PCS | 1.0 (-8.1, 10.1) | 6.2 (-0.4, 12.9) | 10.1 (-7.3, 27.5) |
| PROMIS GH<sup>b</sup> MCS | 3.7 (-5.4, 12.7) | 1.8 (-7.0, 10.6) | 6.1 (-11.4, 23.6) |
| Morphine equivalents<sup>ad</sup> | -2.9 (-7.7, 1.9) | -3.3 (-7.5, 1.0) | -6.7 (-16.4, 3.0) |
| Global perceived effect<sup>b</sup> | 0.2 (-1.4, 1.7) | 0.1 (-1.3, 1.5) | 0.0 (-3.4, 3.5) |
| Satisfaction<sup>b</sup> | 1.3 (-0.5, 3.0) | 1.2 (-0.4, 2.8) | 2.3 (-0.7, 5.4) |
| NRS-UP<sub>(A)</sub><sup>a</sup> | -0.9 (-5.5, 3.7) | -1.4 (-5.4, 2.6) | -2.0 (-14.9, 10.9) |
| BRFSS moderate intensity-equivalent minutes<sup>be</sup> | -637 (-1636, 362) | 572 (-199, 1343) | -82 (-1410, 1245) |

\*Associations are adjusted for age and baseline value of the outcome unless indicated otherwise; models for global perceived effect and satisfaction after treatment did not adjust for baseline value as these outcomes do not apply to baseline (prior to treatment)
<sup>a</sup>Negative values indicate improvement with treatment group as compared to control group
<sup>b</sup>Positive values indicate improvement with treatment group as compared to control group
<sup>c</sup>Change in step counts from baseline to 3 months post-randomization
<sup>d</sup>Morphine equivalent daily dose; because all participants were not taking opioids at baseline, baseline opioid dose was not included as a covariate,
<sup>e</sup>Moderate intensity-equivalent minutes per week
LRFA: lumbar radiofrequency ablation, CI: confidence interval, RMDQ: Roland-Morris Disability Questionnaire NRS: Numerical rating scale, PROMIS GH: Patient-Reported Outcomes Measurement Information System Global Health, PCS: Physical component scale, MCS: Mental component scale, NRS-UP<sub>(A)</sub>: Numerical rating scale of underlying pain without analgesic use, BRFSS: Behavioral risk factor surveillance system

### Primary Outcome (Roland-Morris Disability Questionnaire [RMDQ]) at 3 months post-randomization

Treatment effects on the 3-month RMDQ score primary endpoint were estimated (Table 2) for individual treatment groups (LRFA vs. simulated LRFA; AcTIVE-CBT vs. TBSCE) as well as combined treatments (LRFA/AcTIVE-CBT vs. simulated LRFA/TBSCE). The LRFA treatment effect point estimate favored a beneficial effect of LRFA of moderate magnitude that was not statistically significant (−3.25; 95% CI: -10.18, 3.67). The AcTIVE-CBT point estimate favored a beneficial effect of AcTIVE-CBT. This effect was large and statistically significant (−6.29, 95% CI: -10.97, -1.60). The combined point estimate for LRFA/AcTIVE-CBT favored a beneficial effect of LRFA/AcTIVE-CBT, of large magnitude, that was not statistically significant (−8.37; 95% CI: -21.47, 4.74).

### Secondary Outcomes

Treatment effect point estimates for secondary outcomes showed modest between-group differences and no statistically significant associations were seen for any secondary outcome (Table 2). While not statistically significant, point estimates suggested small beneficial effects of LRFA as compared to simulated LRFA on 3-month NRS (−1.02 NRS points; 95% CI: -5.16, 3.11) and patient satisfaction (1.3 points on a 5-point scale; 95% CI: -0.5, 3.0), alongside non-significant detrimental effects on average daily step counts (6059 fewer steps per day; 95% CI: -12467, 349) and self-reported moderate intensity-equivalent minutes per week (637 fewer minutes per week; 95% CI: -1636, 362). There were no statistically significant effects of AcTIVE-CBT (vs. TBSCE) for any secondary outcome, but treatment effect point estimates favored beneficial effects of AcTIVE-CBT on 3-month PROMIS physical health (6.2 points; 95% CI: -0.4, 12.9) and patient satisfaction (1.2 points; 95% CI: -0.4, 2.8). Additionally, although were no statistically significant effects of combined LRFA/AcTIVE-CBT (vs simulated LRFA/TBSCE) for any secondary outcome, treatment effect point estimates favored beneficial effects of LRFA/AcTIVE-CBT on 3-month PROMIS physical health (10.1 points; 95% CI: -7.3, 27.5) and patient satisfaction (2.3 points; 95% CI: -0.4, 2.8). Generally, treatment effect point estimates for combined LRFA and AcTIVE-CBT treatments suggested additive effects for most secondary outcomes.

### Blinding

Among the 7 participants who were randomized to receive LRFA, 3 guessed that they had received LRFA, 3 guessed that they had received simulated LRFA, and 1 had “no idea”, suggesting randomness in participants’ guesses about their study-group assignments at the 1-month primary endpoint among those receiving LRFA (blinding index 0, 95% CI -0.56 to 0.56). Among the 6 participants who were randomized to receive simulated LRFA, 3 guessed that they had received simulated LRFA, none guessed that they had received LRFA, and 3 had “no idea”, indicating that participants receiving simulated LRFA had a significant tendency to guess that they had received simulated LRFA beyond that expected by chance (blinding index 0.50, 95% CI 0.10 to 0.90). All 3 participants who guessed that they had received simulated LRFA reported that their guess was informed by their impressions of change in pain since the procedure.

### Adverse Events

Eighteen adverse events (AEs) were reported during follow-up (Supplemental Table S3). Five participants reported 1 or more AE, and the remaining 8 participants did not report an AE. Of 18 total AEs reported, 16 (89%) were determined to be unrelated or unlikely to be related to study procedures; examples of unrelated AEs included a cracked tooth and a hearing aid malfunction. One AE determined to be possibly related to the study was a change in leg pain that happened after the lumbar procedure, and another AE determined to be probably related to the study was back stiffness that happened after the lumbar procedure. No SAEs were reported. Some participants reported more than one AE. There were no significant differences in the number of participants reporting AEs between the LRFA and simulated LRFA arms (4 vs. 1, respectively; p=0.27) or AcTIVE-CBT and TBSCE (3 vs. 2, respectively; p=1.00) arms.

## Discussion

This pilot study found that it was feasible to conduct an RCT combining two disparate treatments for CLBP, interventional procedural treatments (LRFA vs. simulated LRFA control) and behavioral pain treatments (AcTIVE-CBT vs. TBSCE control). More than 60% of eligible patients enrolled in the study, more than 80% of those enrolled were randomized, and all randomized received their allocated treatment. Additionally, all randomized participants completed study assessments at 3 months post-randomization. The effects of interventional procedural and behavioral pain treatments on back-related functional limitations appeared to be additive or weakly sub-additive, without suggestion of major synergy or antagonism. While no statistically significant main effects of these treatments on back-related functional limitations were expected due to the pilot’s small sample size, the point estimate for the treatment effect of LRFA vs. simulated LRFA might suggest a beneficial effect of LRFA that was not statistically significant (−3.3 RMDQ points at 3-months post-randomization; 95% CI -10.2, 3.7). Surprisingly, a statistically significant and beneficial treatment effect of AcTIVE-CBT (vs. TBSCE) was found (−6.3 RMDQ points at 3-months post-randomization; 95% CI -11.0, -1.6).

Though not statistically significant, the point estimate for the treatment effect of LRFA vs. simulated LRFA on back-related functional limitations in the current study (3.3 RMDQ points) exceeds the minimum clinically relevant *between-group* effect sizes used for powering large-scale studies, which are 2.25 to 2.5 RMDQ points.^41,42^ However, the 95% CI for the LRFA treatment cannot exclude a large beneficial effect of LRFA (up to 10.2 RMDQ points) or a detrimental effect (up to 3.7 RMDQ points). These results are consistent with prior LRFA trials, which have shown >10% improvement in functional limitations with LRFA at ≥3-month follow-up in two trials[27; 35] but no significant benefit in functional limitations at ≥3-month follow-up in the largest RCT of LRFA to date. [19] Clinical experts in LRFA have noted that prior RCTs showing no meaningful effects may be explained by (1) poor LRFA technique insufficient to adequately lesion the medial branches and (2) suboptimal clinical selection criteria for identifying LRFA candidates.[25] In the current study, we took several steps to optimize medial branch lesioning (parallel electrode placement, large electrodes, and 2 lesions per nerve) and clinical selection (requiring 2 sets of MBBs, using ≤0.5cc anesthetic in MBBs, etc.). These steps may have contributed to the beneficial, albeit non-significant, effect of LRFA in the current pilot trial.

The effect of CBT for pain as estimated in RCTs is already well-known.[37] Nevertheless, the statistically significant, large-magnitude treatment effect of AcTIVE-CBT vs. TBSCE in the current study was larger than we had expected. Standardized mean differences for the immediate effect of CBT on functional limitations from meta-analyses are 0.32 (vs. usual care) and 0.12 (vs. active control),[37] which correspond to treatment effects on the RMDQ scale of approximately 4.0 and 1.5 points, respectively; the latter estimate is substantially smaller than the 6.3-RMDQ-point mean benefit of AcTIVE-CBT vs. TBSCE (an active control) in the current study. This unexpectedly large benefit may be explained by imprecision due to low sample size and the “winner’s curse”[36], in which the size of treatment effects in single RCTs may be inflated, particularly when sample size is low.[17; 34] Another explanation for this may be that despite the fact that participants were not specifically informed which of the AcTIVE-CBT vs. TBSCE treatments was the CBT intervention of interest, the TBSCE control treatment was a self-directed treatment with many fewer sessions than AcTIVE-CBT; due to this, TBSCE may have been perceived by participants as a less effective treatment, leading to a larger estimated treatment effect more comparable to that estimated from an RCT with a usual care comparator arm. Alternatively, the large treatment effect of AcTIVE-CBT on functional limitations in the current study may be due to distinguishing features of the treatment, such as the tailored content from the MAP trial which was specific to CLBP,[8; 9] video-based modality, integration of Fitbit activity monitors, one-on-one (as opposed to group-based) care delivery, or other aspects.

With regards to secondary outcomes, as expected due to the small sample size, estimated treatment effects over control for LRFA and AcTIVE-CBT showed wide 95% Cis, encompassing beneficial, null, or detrimental effects of LRFA. Overall, there was no clear tendency towards a pattern of improvements in the point estimates for the main effects of LRFA or AcTIVE-CBT across secondary outcomes.

To our knowledge, this was the first factorial RCT to combine procedural and psychological treatments for CLBP. It is also the first trial to show the feasibility of recruiting patients with CLBP at the point in usual clinical care when they were already seeking interventional procedural treatments for CLBP (a biomedical approach focused on targeting nociceptive pain generators), yet randomizing them to receive psychologically-oriented behavioral pain treatments, a fundamentally different type of treatment focused on addressing psychological and social components of the biopsychosocial model. Nevertheless, half of eligible participants agreed to participate in the study and were randomized. Despite our study’s triple-blind design with regards to the randomized procedural treatments, the recruitment setting in an interventional pain clinic might have been expected to bias treatment effects in favor of finding relatively larger treatment effects for LRFA than for the active psychological treatment, AcTIVE-CBT. Yet, on the contrary, the opposite pattern was seen. As patients and providers cannot be blinded to psychological treatments, a single-blind design was required with regards to these treatments, and this less stringent blinding may partially explain the large-magnitude treatment effects seen for AcTIVE-CBT.

Our assessments of post-procedural blinding at 1 month after randomization suggested that participants receiving LRFA were unable to guess the treatment they had received, but that participants receiving simulated LRFA were able to guess the treatment they had received. This was surprising, in light of anecdotal observations among blinded research staff who interacted with participants immediately after their procedure on the day of randomization, who noted that participants often spontaneously offered opinions that they had received LRFA (the active treatment) or they could not tell what procedure they had received, but none reported impressions that they had received simulated LRFA. However, no systematic assessment of blinding success was made following the procedure on the day of randomization. This was a potential design flaw, as systematic assessment of blinding at 1 month post-randomization may be affected by participants’ perceptions of their improvement, creating a situation in which those with larger improvements infer they have received the active treatment, and those without improvement infer they have received the control treatment. In other words, blinding assessments may be confounded by participants’ impressions of improvement or lack thereof. This is of particular concern given that improvement in the RMDQ is a strong mediator of favorable patient perceptions after lumbar spinal procedures,[33] and the greatest within-group changes in the RMDQ in the current study occurred *prior* to the assessment of blinding at 1 month post-randomization (Supplemental Figure S2). In future trials of LRFA, the primary blinding assessment should likely take place on the day of the procedure, after the procedure has taken place.

This pilot also had other limitations. Adherence to the randomized procedural treatments was imperfect, with 1 participant randomized to LRFA who had bilateral low back pain yet received LRFA on only one of the 2 painful sides of their low back; given the small sample size, this incomplete adherence might have biased procedural treatment effect estimates towards the null. This occurrence prompted alterations to our study processes that prevented subsequent instances of the same type of nonadherence, and could be used in a future multicenter RCT. Another potential limitation of the study is that the findings with regards to feasibility and effectiveness may not generalize to settings outside the VA health care system, as patients in VA settings generally have higher levels of disability and psychological comorbidity than in civilian health care.[21] We have recently shown that procedural treatment effects on disability outcomes in CLBP can be larger in subgroups of people with higher predicted levels of disability,[23] leaving open the possibility that treatment effects in non-VA settings may be smaller than those in the current study. A specific limitation related to activity tracker-evaluated step counts was that post-MBB improvements in step counts were noted for many participants as compared to their pre-MBB steps, and these improvements had not returned to pre-MBB levels for some participants prior to randomization (even though pain intensity levels had returned to an intensity sufficient to warrant LRFA and were ≥4 on the NRS scale on the day of randomization). This may have affected the results of analyses of step count outcomes. Another limitation is that only male-identifying individuals joined this study; this reflects that the population of older Veterans is predominantly male.

In conclusion, it was feasible to conduct a randomized controlled trial combining procedural and behavioral treatments for CLBP. Significant and large treatment effects of video-based CBT were found, despite that participants were identified at the point of seeking interventional procedures, a fundamentally different type of treatment. Future large-scale factorial trials combining procedural and behavioral treatments should be conducted.

## Supporting information

SELECT Supplemental Tables and Figures

## Acknowledgements

This research was supported by the Department of Veterans Affairs (VA), Veterans Health Administration, Office of Research and Development (ORD), Rehabilitation Research and Development (RR&D) Service under award number I21RX002891. Ms. Tanus, Dr. Nishio, Dr. Williams, Dr. Anderson, Dr. Bambara, Dr. Dawson, Dr. Hsu, Dr. Kim, Dr. Krashin, Dr. Korpak, Mr. Timmons, and Dr. Suri are employees of the VA Puget Sound Health Care System in Seattle, Washington. Drs. Soares and Del Piero were employees at VAPSHCS when this study was conducted. Methodologic consultation was provided by the University of Washington Clinical Learning, Evidence And Research (CLEAR) Center for Musculoskeletal Research Methodologic Core; the CLEAR Center is a Core Center for Clinical Research (CCCR) funded by the National Institute of Arthritis and Musculoskeletal and Skin Diseases (NIAMS) of the National Institutes of Health (NIH) under Award Number P30AR072572. The content is solely the responsibility of the authors and does not reflect the views or opinions of the VA or NIH. The authors have no conflicts of interest.

## DATA AVAILABILITY

Compliant with VA and government regulations at the time of data requests, including the use of VA-level resources for secure electronic access required at the time of, we will use those resources to make de-identified, anonymized datasets available to approved applicants under a formal written agreement. Only completely deidentified/anonymized versions of datasets would be shared. Sharing would only take place under a written agreement prohibiting the recipient from identifying or re-identifying (or taking steps to identify or re-identify) any individual whose data are included in the dataset, and would require a properly submitted Freedom of Information Act request.

## REFERENCES

[1] Altman DG, Royston P. The cost of dichotomising continuous variables. BMJ (Clinical research ed 2006;332(7549):1080.

[2] Bang H, Ni L, Davis CE. Assessment of blinding in clinical trials. Control Clin Trials 2004;25(2):143–156.

[3] Bogduk N. Lumbar Medial Branch Blocks. Practice Guidelines for Spinal Diagnostic and Treatment Procedures, Second Edition. San Francisco: International Spine Intervention Society, 2013. pp. 559-600.

[4] Bogduk N. Lumbar Medial Branch Thermal Radiofrequency Neurotomy. Practice Guidelines for Spinal Diagnostic and Treatment Procedures, Second Edition. San Francisco: International Spine Intervention Society, 2013. pp. 601-630.

[5] Brown DR, Carroll DD, Workman LM, Carlson SA, Brown DW. Physical activity and health-related quality of life: US adults with and without limitations. Quality of life research : an international journal of quality of life aspects of treatment, care and rehabilitation 2014;23(10):2673–2680.

[6] Callahan CM, Unverzagt FW, Hui SL, Perkins AJ, Hendrie HC. Six-item screener to identify cognitive impairment among potential subjects for clinical research. Med Care 2002;40(9):771–781.

[7] Caudill MA. Managing Pain Before It Manages You: The Guilford Press, 2016.

[8] Cherkin DC, Anderson ML, Sherman KJ, Balderson BH, Cook AJ, Hansen KE, Turner JA. Two-Year Follow-up of a Randomized Clinical Trial of Mindfulness-Based Stress Reduction vs Cognitive Behavioral Therapy or Usual Care for Chronic Low Back Pain. JAMA 2017;317(6):642–644.

[9] Cherkin DC, Sherman KJ, Balderson BH, Cook AJ, Anderson ML, Hawkes RJ, Hansen KE, Turner JA. Effect of Mindfulness-Based Stress Reduction vs Cognitive Behavioral Therapy or Usual Care on Back Pain and Functional Limitations in Adults With Chronic Low Back Pain: A Randomized Clinical Trial. JAMA 2016;315(12):1240–1249.

[10] Chiarotto A, Boers M, Deyo RA, Buchbinder R, Corbin TP, Costa LOP, Foster NE, Grotle M, Koes BW, Kovacs FM, Lin CC, Maher CG, Pearson AM, Peul WC, Schoene ML, Turk DC, van Tulder MW, Terwee CB, Ostelo RW. Core outcome measurement instruments for clinical trials in nonspecific low back pain. Pain 2018;159(3):481–495.

[11] Chou R, Deyo R, Friedly J, Skelly A, Hashimoto R, Weimer M, Fu R, Dana T, Kraegel P, Griffin J, Grusing S, Brodt E. Noninvasive Treatments for Low Back Pain [Internet]. Noninvasive Treatments for Low Back Pain. Rockville (MD), 2016.

[12] Deyo RA, Dworkin SF, Amtmann D, Andersson G, Borenstein D, Carragee E, Carrino J, Chou R, Cook K, Delitto A, Goertz C, Khalsa P, Loeser J, Mackey S, Panagis J, Rainville J, Tosteson T, Turk D, Von Korff M, Weiner DK. Report of the NIH Task Force on research standards for chronic low back pain. Physical therapy 2015;95(2):e1–e18.

[13] Dieleman JL, Cao J, Chapin A, Chen C, Li Z, Liu A, Horst C, Kaldjian A, Matyasz T, Scott KW, Bui AL, Campbell M, Duber HC, Dunn AC, Flaxman AD, Fitzmaurice C, Naghavi M, Sadat N, Shieh P, Squires E, Yeung K, Murray CJL. US Health Care Spending by Payer and Health Condition, 1996-2016. JAMA 2020;323(9):863–884.

[14] Dreyfuss P, Halbrook B, Pauza K, Joshi A, McLarty J, Bogduk N. Efficacy and validity of radiofrequency neurotomy for chronic lumbar zygapophysial joint pain. Spine 2000;25(10):1270–1277.

[15] Gellhorn AC, Katz JN, Suri P. Osteoarthritis of the spine: the facet joints. Nat Rev Rheumatol 2013;9(4):216–224.

[16] Heapy AA, Higgins DM, LaChappelle KM, Kirlin J, Goulet JL, Czlapinski RA, Buta E, Piette JD, Krein SL, Richardson CR, Kerns RD. Cooperative pain education and self-management (COPES): study design and protocol of a randomized non-inferiority trial of an interactive voice response-based self-management intervention for chronic low back pain. BMC Musculoskelet Disord 2016;17:85.

[17] Ioannidis JP. Why Most Discovered True Associations Are Inflated. Epidemiology 2008;19:640–648.

[18] Jensen MP, Turner JA, Romano JM, Fisher LD. Comparative reliability and validity of chronic pain intensity measures. Pain 1999;83(2):157–162.

[19] Juch JNS, Maas ET, Ostelo R, Groeneweg JG, Kallewaard JW, Koes BW, Verhagen AP, van Dongen JM, Huygen F, van Tulder MW. Effect of Radiofrequency Denervation on Pain Intensity Among Patients With Chronic Low Back Pain: The Mint Randomized Clinical Trials. JAMA 2017;318(1):68–81.

[20] Katz JN. Lumbar disc disorders and low-back pain: socioeconomic factors and consequences. J Bone Joint Surg Am 2006;88 Suppl 2:21–24.

[21] Kneeman J, Battalio SL, Korpak A, Cherkin DC, Luo G, Rundell SD, Suri P. Predicting Persistent Disabling Low Back Pain in Veterans Affairs Primary Care Using the STarT Back Tool. PM&R 2021;13(3):241-249.

[22] Lee CH, Chung CK, Kim CH. The efficacy of conventional radiofrequency denervation in patients with chronic low back pain originating from the facet joints: a meta-analysis of randomized controlled trials. Spine J 2017.

[23] Liu P, Wu Y, Xiao Z, Gold LS, Heagerty PJ, Annaswamy T, Friedly J, Turner JA, Jarvik JG, Suri P. Estimating individualized treatment effects using a risk-modeling approach: an application to epidural steroid injections for lumbar spinal stenosis. Pain 2022.

[24] Maas ET, Ostelo RW, Niemisto L, Jousimaa J, Hurri H, Malmivaara A, van Tulder MW. Radiofrequency denervation for chronic low back pain. Cochrane Database Syst Rev 2015(10):CD008572.

[25] McCormick ZL, Vorobeychik Y, Gill JS, Kao MJ, Duszynski B, Smuck M, Stojanovic MP. Guidelines for Composing and Assessing a Paper on the Treatment of Pain: A Practical Application of Evidence-Based Medicine Principles to the Mint Randomized Clinical Trials. Pain medicine (Malden, Mass 2018;19(11):2127–2137.

[26] Montgomery AA, Peters TJ, Little P. Design, analysis and presentation of factorial randomised controlled trials. BMC Med Res Methodol 2003;3:26.

[27] Moussa WM, Khedr W. Percutaneous radiofrequency facet capsule denervation as an alternative target in lumbar facet syndrome. Clin Neurol Neurosurg 2016;150:96–104.

[28] Murray CJ, Atkinson C, Bhalla K, Birbeck G, Burstein R, Chou D, Dellavalle R, Danaei G, Ezzati M, Fahimi A, Flaxman D, Foreman, Gabriel S, Gakidou E, Kassebaum N, Khatibzadeh S, Lim S, Lipshultz SE, London S, Lopez, MacIntyre MF, Mokdad AH, Moran A, Moran AE, Mozaffarian D, Murphy T, Naghavi M, Pope C, Roberts T, Salomon J, Schwebel DC, Shahraz S, Sleet DA, Murray, Abraham J, Ali MK, Bartels DH, Chen H, Criqui MH, Dahodwala, Jarlais, Ding EL, Dorsey ER, Ebel BE, Fahami, Flaxman S, Flaxman AD, Gonzalez-Medina D, Grant B, Hagan H, Hoffman H, Leasher JL, Lin J, Lozano R, Lu Y, Mallinger L, McDermott MM, Micha R, Miller TR, Mokdad AA, Narayan KM, Omer SB, Pelizzari PM, Phillips D, Ranganathan D, Rivara FP, Sampson U, Sanman E, Sapkota A, Sharaz S, Shivakoti R, Singh GM, Singh D, Tavakkoli M, Towbin JA, Wilkinson JD, Zabetian A, Alvardo M, Baddour LM, Benjamin EJ, Bolliger I, Carnahan E, Chugh SS, Cohen A, Colson KE, Cooper LT, Couser W, Dabhadkar KC, Dellavalle RP, Dicker D, Duber H, Engell RE, Felson DT, Finucane MM, Fleming T, Forouzanfar MH, Freedman G, Freeman MK, Gillum RF, Gosselin R, Gutierrez HR, Havmoeller R, Jacobsen KH, James SL, Jasrasaria R, Jayarman S, Johns N, Lan Q, Meltzer M, Mensah GA, Michaud C, Mock C, Moffitt TE, Nelson RG, Olives C, Ortblad K, Ostro B, Raju M, Razavi H, Ritz B, Sacco RL, Shibuya K, Silberberg D, Singh JA, Steenland K, Taylor JA, Thurston GD, Vavilala MS, Vos T, Wagner GR, Weinstock MA, Weisskopf MG, Wulf S. The state of US health, 1990-2010: burden of diseases, injuries, and risk factors. JAMA 2013;310(6):591–608.

[29] Patient-Centered Outcomes Research Institute. Topics in the Prioritization Pathway (Pain), Vol. 2017, 2017.

[30] Paul SS, Tiedemann A, Hassett LM, Ramsay E, Kirkham C, Chagpar S, Sherrington C. Validity of the Fitbit activity tracker for measuring steps in community-dwelling older adults. BMJ Open Sport Exerc Med 2015;1(1):e000013.

[31] Roland M, Fairbank J. The Roland-Morris Disability Questionnaire and the Oswestry Disability Questionnaire. Spine (Phila Pa 1976) 2000;25(24):3115–3124.

[32] Strath SJ, Bassett DR, Jr., Ham SA, Swartz AM. Assessment of physical activity by telephone interview versus objective monitoring. Medicine and science in sports and exercise 2003;35(12):2112–2118.

[33] Suri P, Pashova H, Heagerty PJ, Jarvik JG, Turner JA, Comstock BA, Bauer Z, Annaswamy TM, Nedeljkovic SS, Wasan AD, Friedly JL. Short-term improvements in disability mediate patient satisfaction after epidural corticosteroid injections for symptomatic lumbar spinal stenosis. Spine (Phila Pa 1976) 2015;40(17):1363–1370.

[34] Tugwell P, Knottnerus JA. A statistic to avoid being misled by the “winners curse”. J Clin Epidemiol 2018;103:vi–viii.

[35] van Kleef M, Barendse GA, Kessels A, Voets HM, Weber WE, de Lange S. Randomized trial of radiofrequency lumbar facet denervation for chronic low back pain. Spine 1999;24(18):1937–1942.

[36] van Zwet E, Schwab S, Senn S. The statistical properties of RCTs and a proposal for shrinkage. Stat Med 2021;40(27):6107–6117.

[37] Williams ACC, Fisher E, Hearn L, Eccleston C. Psychological therapies for the management of chronic pain (excluding headache) in adults. Cochrane Database Syst Rev 2020;8:CD007407.

[38] Yore MM, Ham SA, Ainsworth BE, Kruger J, Reis JP, Kohl HW, 3rd, Macera CA. Reliability and validity of the instrument used in BRFSS to assess physical activity. Medicine and science in sports and exercise 2007;39(8):1267–1274.

