## Supplementary material for "Combining Procedural and Behavioral Treatments for Chronic Low Back Pain: A Pilot Feasibility Randomized Controlled Trial": SELECT Supplemental Tables and Figures

| **Supplemental Table S1. Summary of inclusion and exclusion modification during the pilot study** | | |
| --- | --- | --- |
| **Original Criterion** | **Change** | **Justification for Change** |
| Exclusion of insulin-using diabetics (for whom elevated blood glucose may occur) or those with hemoglobin A1C levels ≥ 8.0 | Removed as an exclusion criterion | Initially, we had excluded insulin-using diabetics from study participation because we thought that post-RFA glucose changes due to steroid use in the simulated LRFA group might potentially unblind patients as to their treatment allocation. However, as all the participating providers already routinely used corticosteroids after LRFA to reduce post-RFA ‘neuritis’ flares (short term flares of pain that come directly from the nerve lesioning/burning), and post-LRFA glucose changes were dealt with by routine education directing patients to more closely monitor their sugars post-procedure, there was no potential for differential unblinding between treatment vs. control groups. Because of this, there was no reason to exclude insulin-using diabetics from the study. |
| Exclusion of patients with non-compliance with Fitbit use during run-period. Participants who did not wear the Fitbit unit on at least 3 of 7 days during the test period, for at least 6 hours each day, were considered non-compliant. | Removed as an exclusion criterion | It was felt that our requirements for Fitbit use/compliance during the run-in period were overly demanding for participants unfamiliar with such technology, not providing sufficient time for participants to adapt to using such technology. Moreover, it was felt that the run-in period might exclude patients who might otherwise benefit from Fitbit use (and have the capacity to improve their ability to use the Fitbit and thus begin to benefit from it) over the course of the study. |
| Inclusion criterion of “positive responses” to lumbar MBBs defined as ≥50% pain improvement of typical low back pain and anesthetic type-concordant duration of pain relief (onset of typical lumbar back pain relief within 30 mins, relief lasting at least 30 mins after the onset of initial pain relief, and duration of relief of typical lumbar back pain concordant with that expected for anesthetic type [<12 hours for lidocaine, <36 hours for bupivicaine]). | This criterion was altered to allow participants with ≥50% relief with 2 or more sets of medial branch blocks to participate in the study, instead of ≥80% relief with 2 or more sets of blocks; to remove the requirement that pain improve with an activity that could be performed in the fluoroscopy suite; and to remove the requirement that the duration of relief of typical lumbar back pain be concordant with that expected for anesthetic type [<12 hours for lidocaine, <36 hours for bupivicaine]).  The revised criterion for “positive responses” to lumbar MBBs was:  1) ≥50% pain improvement of typical low back pain, and 2) onset of typical lumbar back pain relief within 30 mins of MBBs, and relief lasting at least 30 mins after the onset of initial pain relief after MBBs | The original criterion was felt to be inappropriately restrictive so as not to be representative of typical clinical practice in our facility and the VA more broadly. Changes were made to enhance the generalizability of the study population and study results to reflect the typical standards used in the VA. Also, clinical LRFA characteristics in our facility suggested that the pilot would be unlikely to recruit sufficient study participants if defining positive responses using the initial definition |
| Any prior lumbar RFA or prior lumbar spine surgery | This criterion was made more specific and split into two separate criteria: (1) prior lumbar spine surgery involving the levels where LRFA was to be performed within the past 2 years; (b) ever having received lumbar fusion involving the levels where LRFA was to be performed | Requiring participants to have had no prior lumbar spine surgery was determined to be overly restrictive and not reflective of standard clinical practice, where LRFA may often by done for patients who have previously had lumbar spine surgery. |
| Excessive alcohol consumption or drug use as determined by the TICS questionnaire (1+ positive answer) | Removed this as an exclusion criterion | The TICS criteria was previously inadvertently included as a study exclusion. On reviewing our forms, we determined the TICS exclusion inapplicable to the current study population, and likely to exclude many Veterans who are appropriate for the study-related treatments and who benefit from these treatments in usual clinical care. |
| Radiating pain, numbness, or tingling below the level of the knee attributed to a spinal source, and not explained by other conditions | This criterion was changed to:  Clinical suspicion of CLBP due to specific lumbar spine-related syndromes including lumbosacral radicular syndrome (radiculopathy), symptomatic lumbar spinal stenosis (neurogenic claudication), with confirmatory imaging findings, spinal instability requiring surgery, or other “red flag” conditions (infection/ malignancy/ fracture) | The initial criterion was felt to be overly broad and too ambiguous for participating providers attempting to ascertain preliminary eligibility. |
| Low back pain intensity numerical rating scale (NRS) > 4 (must be 5 or higher) | The minimum NRS pain intensity threshold needed to participate in the study was reduced from 5/10 to 4/10, and was modified to account for pain that may be associated with (and worsened by) basic activities. The criterion was changed to:  Low back pain intensity numerical rating scale (NRS) ≥ 4 (must be 4 or higher) | A minimum pain intensity required for participation of 4/10 is a more common standard used for pain studies and a value that is accepted as corresponding to “moderate” pain intensity. |
| Prior CBT for chronic pain | This exclusion criterion was made more specific. Prior CBT was defined as a full course of CBT focused on pain, ≥4 sessions. | The initial criterion was felt to be overly broad and might exclude participants who had one session of CBT only, or a session of another treatment that included a small amount of CBT, or CBT for another condition that was not focused on pain. |
| Moderate or severe limitations in ambulation due to conditions other than CLBP | Removed the exclusion criteria of moderate or severe ambulatory limitations | The exclusion of moderate or severe ambulatory limitations limited generalizability to those with reasonable mobility, and these individuals can still benefit from treatments for pain. Thus, this exclusion was removed. |

| **Supplemental Table S2. Behavioral pain treatments provided to study participants*** | | |
| --- | --- | --- |
|  | **AcTIVE-CBT**  See full treatment manual provided as supplemental file | **TBSCE** |
| General descriptions and content prior to first session with psychologist | Eight 60-minute sessions conducted by video telehealth and psychologist-delivered.  Participant will receive basic instruction on Fitbit Zip use from research staff prior to randomization.  Participant will receive ongoing research staff support regarding Fitbit use post-randomization, including telephone contacts by research staff at approximately 2 weeks, 6 weeks, and 10 weeks post-randomization.  Research staff will produce regular reports on participant Fitbit output and compliance prior to each treatment session, which will be made available to the psychologist and the data relayed to participants during the treatment sessions. | Telephone, based, self-guided bibliography treatment with 1 hour introductory session and one booster session with a psychologist.  TBSCE is an active yet lower-dose intensity treatment as compared to AcTIVE-CBT. TBSCE will include pain education and CBT-related education and guidance through Dr. Margaret Caudill’s CBT ‘***Managing Chronic Pain***’ workbook (referred to as ‘‘*Caudill’* below)  Participant will receive basic instruction on Fitbit Zip use from research staff prior to randomization.  Participant will receive ongoing research staff support regarding Fitbit use post-randomization including telephone contacts by research staff at approximately 2 weeks, 6 weeks, and 10 weeks post-randomization. |
| Session 1 | **60 min. session by rehabilitation psychologist** Welcome and Introductions; Pain and the Brain | **60 min. telephone session by rehabilitation psychologist**  Welcome and Introduction; Introduction to Materials (TBSCE Manual and ‘Managing Pain Before It Manages You’ workbook by Caudill); Overview of the Treatment Structure; Questions  Reading Assignments   - Chapter 1 (Beginning to Take Control of Your Pain) - Chapter 2 (Understanding Pain)   Topics:   - Education about different types of pain - Self-assessment about how you currently cope with pain. |
| Session 2 | **60 min. session by rehabilitation psychologist.**  Getting Active! Goal-Setting, Pacing, and Managing Flare-Ups | Reading Assignments   - Chapter 3 (The Mind-Body Connection)   Topics:   - Pain as a form of Chronic Stress - Relaxation Response |
| Session 3 | **60 min. session by rehabilitation psychologist.**  Thoughts, Feelings, and Pain | Reading Assignments   - Chapter 4 (The Body-Mind Connection)   Topics:   - Increasing activities - How doing activity improves your mood - Pleasant activities |
| Session 4 | **60 min. session by rehabilitation psychologist.**  Challenging Automatic Thoughts: Part I | Reading Assignments:   - Chapter 5 (The Power of the Mind, pages 97-112)   Topics:   - How to use powerful cognitive techniques to change your mood. - Recognize your “self-talk”, or “automatic thoughts” - Recognize thought distortions |
| Session 5 | **60 min. session by rehabilitation psychologist.**  Challenging Automatic Thoughts: Part II | Reading Assignments:  Chapter 5 (The Power of the Mind).  Topics:   - How to use powerful cognitive techniques to change your mood. - Recognize your “self-talk”, or “automatic thoughts” - Recognize thought distortions   **10-20 min psychologist contact by phone conducted between 1-2 months after the initial TBSCE session, to encourage progress with written materials and self-directed progression.** |
| Session 6 | **60 min. session by rehabilitation psychologist.**  Thinking about Thoughts: Review and Trouble-Shooting | Reading Assignments:  Chapter 6 (Adopting Healthy Attitudes)  Topics:   - Health attitudes |
| Session 7 | **60 min. session by rehabilitation psychologist.**  Pain Beliefs and Behaviors: Skill Review and Sleep | Reading Assignments:  Chapter 8 (Effective Communication)  Topics:   - Assertiveness - Active Listening |
| Session 8 | **60 min. session by rehabilitation psychologist.**  Maintaining Gains and Coping with Setbacks | Reading Assignments:   - Chapter 9 (Effective Problem Solving) - Chapter 10 (The End of The Beginning)   Topics:   - Setting Goals - Applying your coping skills to problems - Relapse Prevention - Coping with Pain During Flare-Ups |
| *See the AcTIVE-CBT manual for details of treatment. | | |

| **Supplemental Table S3. Number of Participants with Adverse Events*** | | | | | |
| --- | --- | --- | --- | --- | --- |
|  | **Procedural Interventions** | | **Behavioral Interventions** | | **Total** |
|  | **LRFA** | **Simulated LRFA** | **AcTIVE-CBT** | **TBSCE** |  |
| **Surgery** | 1 | 0 | 1 | 0 | 1 |
| **ED/Urgent Care** | 3 | 1 | 2 | 2 | 4 |
| **Hospitalization** | 0 | 0 | 0 | 0 | 0 |
| **ICU** | 0 | 0 | 0 | 0 | 0 |
| **Allergic Reaction** | 2 | 0 | 2 | 0 | 2 |
| **Other Complications** | 5 | 1 | 6 | 0 | 6 |
| **Total AEs** | 11* | 2 | 11 | 2 | 13 |

*There were 18 total adverse events (AEs) reported during the study, with 0 serious AEs. **14** AEs were classified as unrelated, **2** unlikely, **1** possibly related, **1** probably related, and **0** definitely related. One participant reported 7 AEs (4 unrelated, 2 unlikely, 1 probably related).

**Supplemental Table S4. Number of participants by AE occurrence (Procedural Interventions)***

|  | **LRFA** | **Simulated LRFA** | **Total** |
| --- | --- | --- | --- |
| **AEs** | 4 | 1 | 5 |
| **No AEs** | 3 | 5 | 8 |
| **Total** | 7 | 6 | 13 |

**P*-value from Fisher’s Exact Test: 0.27

**Supplemental Table S5. Number of participants by AE occurrence (Behavioral Interventions)***

|  | **AcTIVE-CBT** | **TBSCE** | **Total** |
| --- | --- | --- | --- |
| **AEs** | 3 | 2 | 5 |
| **No AEs** | 4 | 4 | 8 |
| **Total** | 7 | 6 | 13 |

**P*-value from Fisher’s Exact Test: 1.00

**Supplemental Figure S1.** Participant flow through study processes.

**Supplemental Figure S2.** Plot of mean RMDQ values by visit and procedural treatment group*

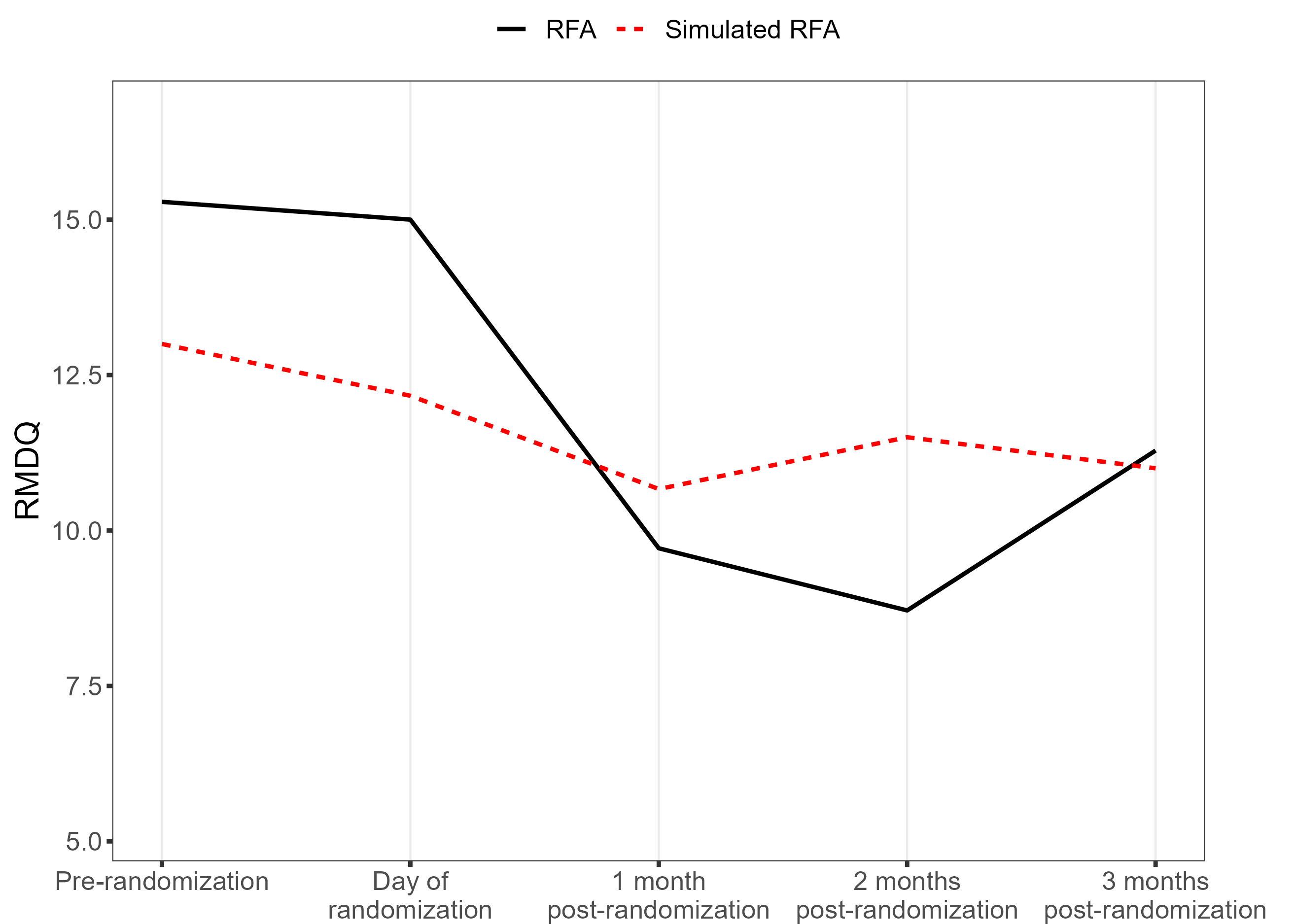

*Pre-randomization (following 2^nd^ set of MBBs, prior to day of randomization), Day of randomization (pre-procedure).

**Supplemental Figure S3.** Plot of mean RMDQ values by visit and behavioral treatment group*

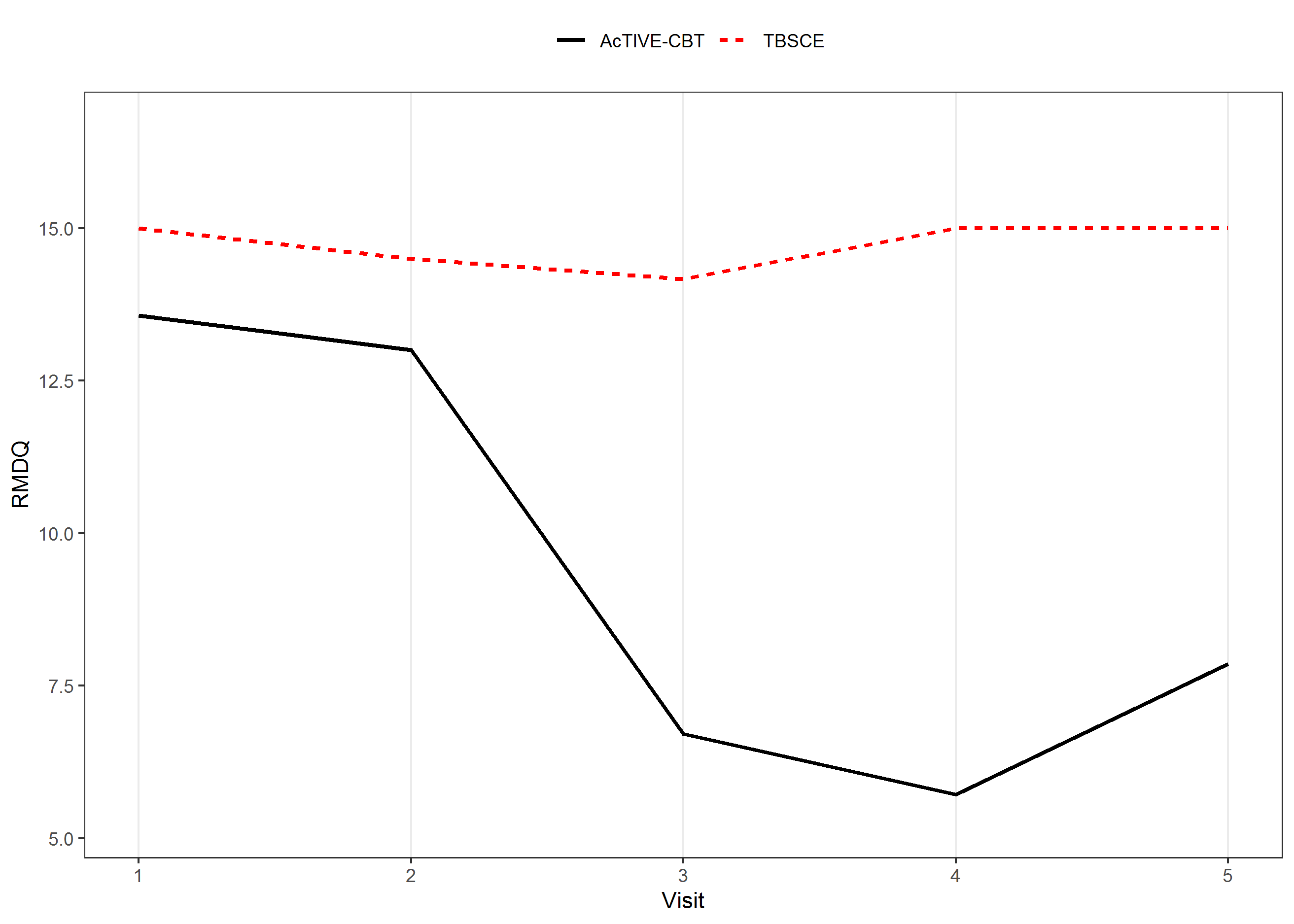

*Pre-randomization (following 2^nd^ set of MBBs, prior to day of randomization), Day of randomization (pre-procedure).

**Supplemental Figure S4.** Plot of mean RMDQ values by visit and treatment group*

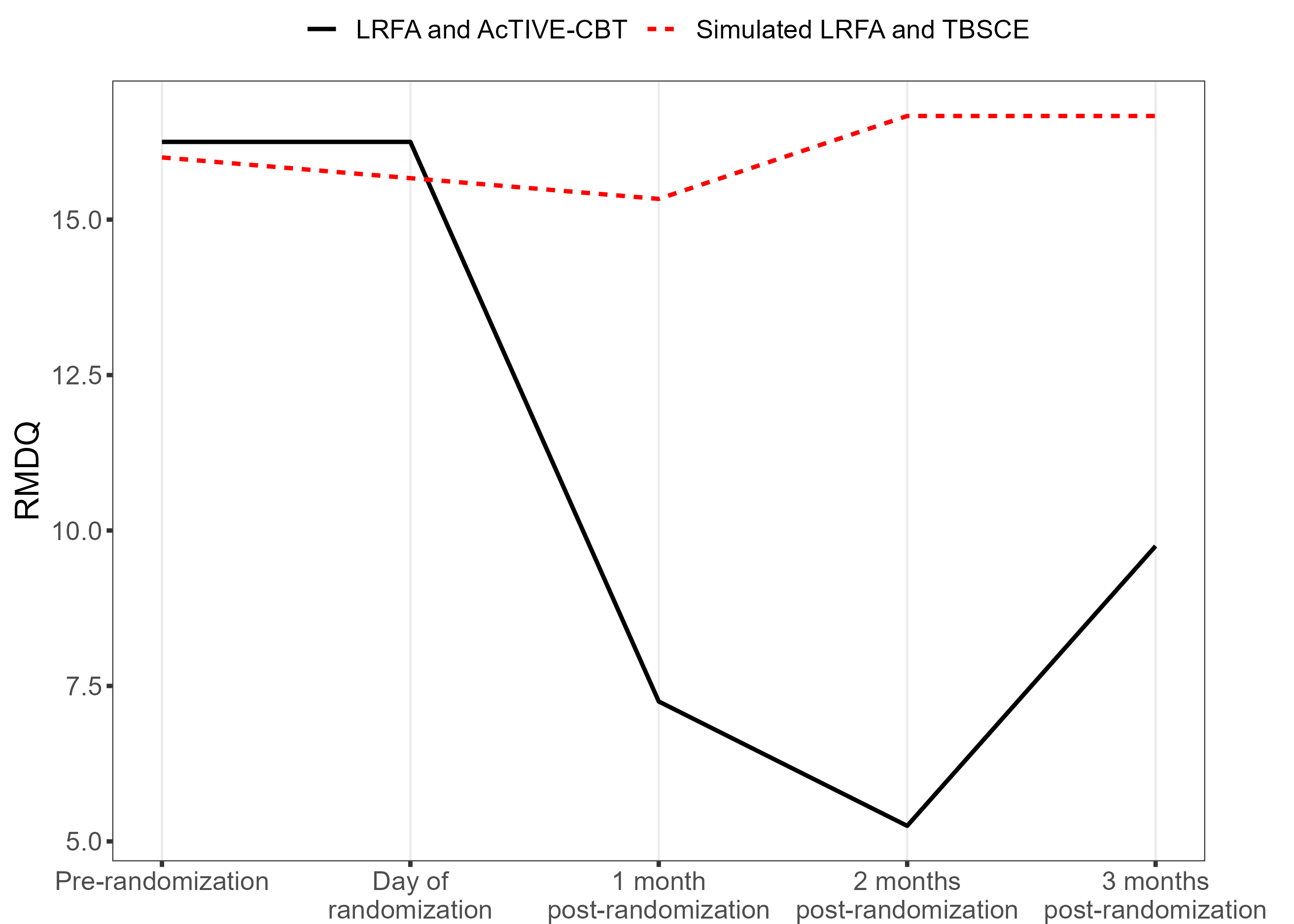

*Pre-randomization (following 2^nd^ set of MBBs, prior to day of randomization), Day of randomization (pre-procedure).
